# The Association of NICU Strain with Neonatal Mortality and Morbidity

**DOI:** 10.1101/2024.07.07.24310050

**Authors:** Elizabeth G. Salazar, Molly Passarella, Brielle Formanowski, Jeannette Rogowski, Erika Edwards, Ciaran Phibbs, Scott A. Lorch

## Abstract

**Objective:** To examine the association of admission NICU strain with neonatal mortality and morbidity.

**Study Design:** 2008-2021 South Carolina cohort using linked vital statistics and discharge data of 22-44 weeks GA infants, born at hospitals with ≥ level 2 unit and ≥5 births of infants <34 weeks GA/year. The exposure was tertiles of admission NICU strain, defined as the sum of infants <44 weeks GA with a congenital anomaly plus all infants born <33 weeks GA at midnight on the day of birth. We used Poisson generalized linear mixed models to examine the association of exposure to strain with the primary outcome of a composite of mortality and term and preterm morbidities adjusting for patient and hospital characteristics.

**Results:** We studied 64,647 infants from 30 hospitals. High strain was associated with increased risk of mortality and morbidity adjusting for patient/hospital factors (aIRR 1.07, 95% CI 1.01 – 1.12).

**Conclusion:** NICU strain is associated with increased adverse outcomes.

## Introduction

Annually in the United States, 12% of infants are admitted to the neonatal intensive care unit (NICU), with substantial variation in outcomes.^1^ Hospital preterm mortality rates vary by 15-fold and rates of morbidities, such as chronic lung disease (CLD) and severe intraventricular hemorrhage (sIVH), vary by 1- to 3-fold.^2–4^ Persistent hospital outcome variation while controlling for patient acuity suggests that hospital factors play a key role in this variation.^4,5^ While availability of hospital resources, measured through AAP neonatal levels of care, and experience with the neonatal patient population, measured by volume, are both associated with neonatal outcomes, ^6–8^ the persistence of inter-hospital variation after accounting for these factors suggests that other hospital measures may contribute to this variation. little research has examined the role of NICU capacity strain on neonatal outcomes despite a robust association between ICU strain and adult outcomes.

One potential factor is ICU strain. ICU strain is the ICU’s time-varying ability to meet care demands with available resources for potential patients.^9^ Strain is influenced by physical and human resources as well as patient volumes, acuity, and specialized needs.^10^ Measures of ICU strain frequently encompass the concepts of patient census, acuity, admissions, discharges, and staffing. The most commonly used measures of ICU strain include ICU census (potentially risk adjusted) and number of admissions.^11,12^ Increased ICU strain on an adult patient’s day of admission is associated with decreased in-hospital survival and safety practices.^11,13–16^ Despite strong evidence in the adult ICU, the role of *NICU* strain as a driver of hospital variation has been understudied. Limited published studies report an association of NICU census with risk of infection in very preterm infants^17^ and census with likelihood of discharge.^18^ By influencing quality of care delivered, NICU strain is hypothesized to ultimately influence patient outcomes (Conceptual Model, Figure 1).

**Figure 1.**
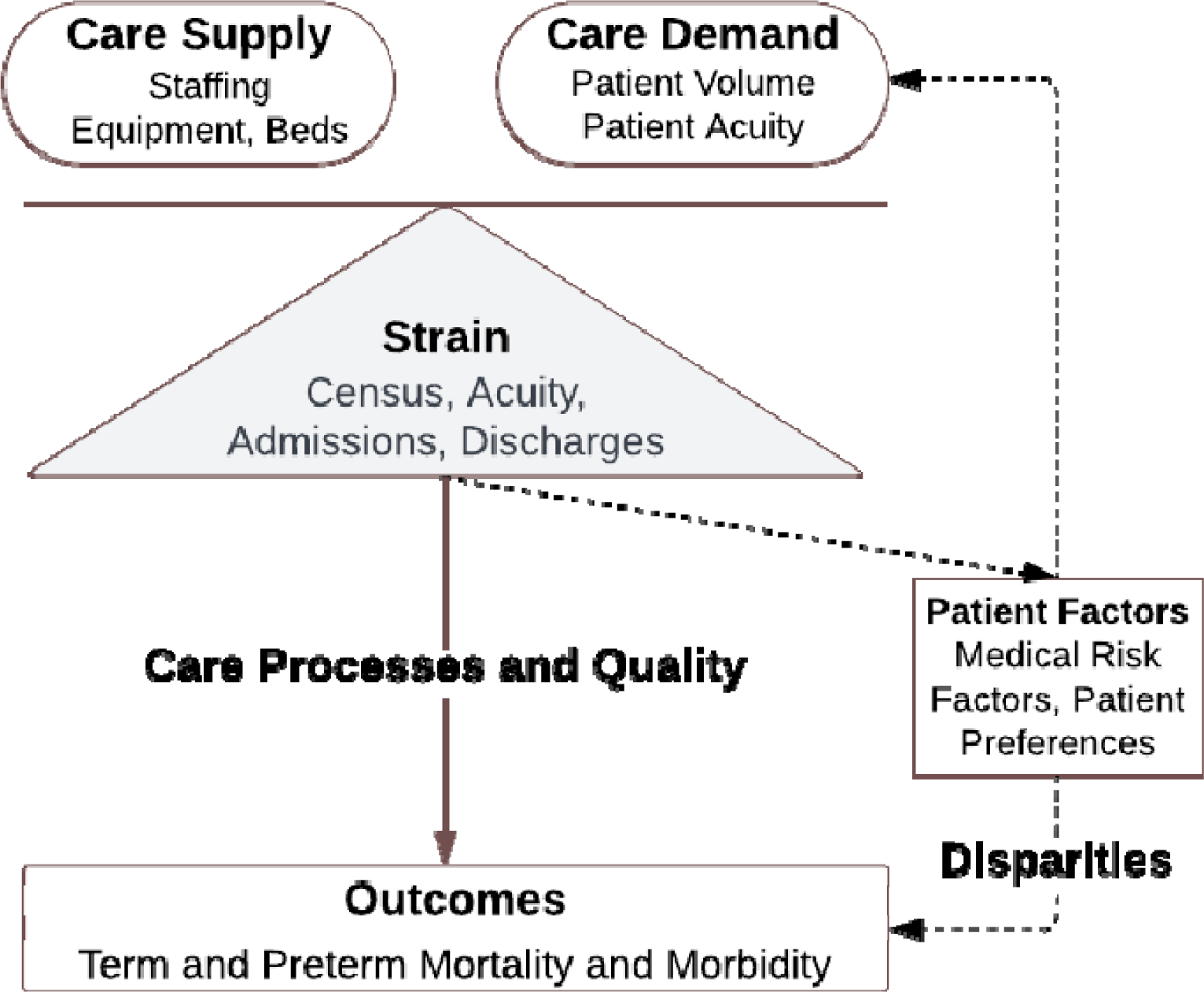
Conceptual Model of NICU Strain and its Relationship with Processes and Outcomes of Care.

Thus, the study objective was to examine the association of NICU strain at admission with in-hospital term and preterm mortality and morbidity. We chose to focus on strain at admission as prior research supports both that delivery hospital has a strong influence on neonatal outcomes^5^ and admission ICU strain is associated with adult mortality.^11^ We used a novel measure of NICU that incorporates concepts of census and patient risk by measuring the standardized daily sum of infants < 44 weeks gestational age (GA) with a congenital anomaly plus all infants born < 33 weeks GA. We hypothesized that increased NICU strain would be associated with worse neonatal outcomes when controlling for both patient-level and hospital-level factors.

## Methods

### Data and Study Population

Using 2008 to 2021 linked South Carolina vital statistics and hospital records, we conducted a retrospective cohort study of infants between born 22 and 44 weeks GA. Infant birth and death certificate data were linked to birthing parent and infant hospital administrative data by the South Carolina Department of Health with a reported birthing parent-infant match rate of 96-99% using described methods.^19^ We used the merged American Hospital Association (AHA) Annual Survey of Hospitals data to obtain hospital characteristics.^20^ Infants were excluded if they had missing birth certificate, birthing parent or infant hospital data or a birth weight greater than five standard deviations from the mean for GA, suggesting that one or both of these variables were miscoded (Supplemental Figure 1).^21^ To restrict the study population to birthing hospitals with NICUs, included infants were born at hospitals with neonatal care level 2 or greater and with ≥ 5 births of infants <34 weeks GA per year (Supplemental Figure 1). Included infants also had a revenue code of level 2 or greater, which is consistent with the receipt of care in a NICU.

This study followed the Strengthening the Reporting of Observational Studies in Epidemiology (STROBE) guidelines.^22^ The Children’s Hospital of Philadelphia institutional review board determined that the abstracted data did not meet the requirements of human subjects research. Data use was approved by the South Carolina Department of Health.

### Study Measures and Variables

The primary exposure of interest was the NICU strain at admission. NICU strain was measured at admission given both the neonatal literature supporting the importance of the delivery hospital for patient outcomes^5^ and the adult literature emphasizing the role of admission strain on outcomes.^11^ Admission NICU strain was defined using a standardized measure of the daily census of higher-risk infants, defined as infants < 44 weeks GA with a congenital anomaly plus all infants born < 33 weeks GA. We included infants <44 weeks GA with a congenital anomaly in our definition as we know these infants are more likely to require resource intensive care, and thus serve as a proxy for unit-level acuity.^23^ We included all infants born < 33 weeks GA because prematurity also places infants at risk for increased resource utilization and potential acuity.^24^ We used a 33 week GA threshold instead of a 34 week GA threshold used to define NICUs as we aimed to identify a patient population at higher risk for resource utilization and acuity. Standardization was performed by subtracting the average annual hospital census of this higher-risk population from this daily census of this higher-risk population and then dividing by the average annual hospital census of this higher-risk population. Standardizing by a unit’s annual daily census of high-risk infants distinguishes this value from volume, as the measure assesses the deviation of the daily census of high-risk infants from the number of such patients that a hospital usually cares for. After creating a separate group of patients born on days of zero strain, NICU strain was then divided into tertiles of low, typical, and high. Strain was assigned based on the value for the day prior to the patient’s birth to ensure the exposure occurred prior to the outcome. We chose a definition of NICU strain that intentionally did not incorporate care resources, such as nursing staffing ratios, as these data are typically unavailable for hospitals on a daily basis and annual values vary by other hospital factors such as financial health and stability.^25^

The primary outcome was a composite of mortality and morbidity. For preterm infants, morbidity included severe (grade 3 or 4) intraventricular hemorrhage (IVH), necrotizing enterocolitis (NEC), surgical retinopathy of prematurity (ROP), chronic lung disease (CLD) or infection as defined by ICD codes (Supplemental Table 1).^26^ For term infants, morbidity included moderate and severe unexpected newborn complications as previously defined.^27^ Outcomes for infants transferred after delivery were assigned to the birth hospital given literature supporting the role of the delivery hospital on neonatal outcomes.^5^

### Covariate Definitions

Patient-level covariates included gestational age (by week), infant sex,^28^ small for gestational age (<10^th^ percentile for GA using Fenton growth chart),^29,30^ multiple gestation,^28^ and congenital anomaly,^23^ which have all been previously associated with the aforementioned outcome measures. Birthing parent covariates included race and ethnicity,^31^ age,^32^ insurance,^33^ education,^34^ smoking,^35^ any diabetes,^36^ any hypertension,^37^ BMI,^38^ mode of delivery,^39^ and birth year^40^ given known association with outcomes. We also examined hospital characteristics including ownership,^41^ rurality,^41^ number of NICU beds,^42^ NICU level of care,^43^ and annual birth volume^6^ (Supplemental Table 2).

### Statistical Analyses

Descriptive statistics were reported using counts and percentages for categorical variables and means and SDs for continuous variables. We evaluated associations using χ2 tests for dichotomous variables and analysis of variance for continuous variables. We examined the hospital level variability of patient level NICU strain using boxplots (Figure 2). We examined the overall association of tertiles of NICU strain with mortality and morbidity by using multivariable Poisson generalized linear mixed models with adjustment for birthing parent and infant to estimate risk ratios (Figures 3 and 4). After exploring described hospitals factors, we chose to use a fixed effect for hospital which captured the variation associated with these variables. We also used a fixed effect for hospital with neonatal level of care to isolate the effect of level of care while also addressing other hospital specific variation. Inclusion of this hospital fixed effect allows for one to interpret the strain variable in these models as the effect of changes in strain within a given hospital on outcomes. Analyses were performed using Stata, version 18.

**Figure 2.**
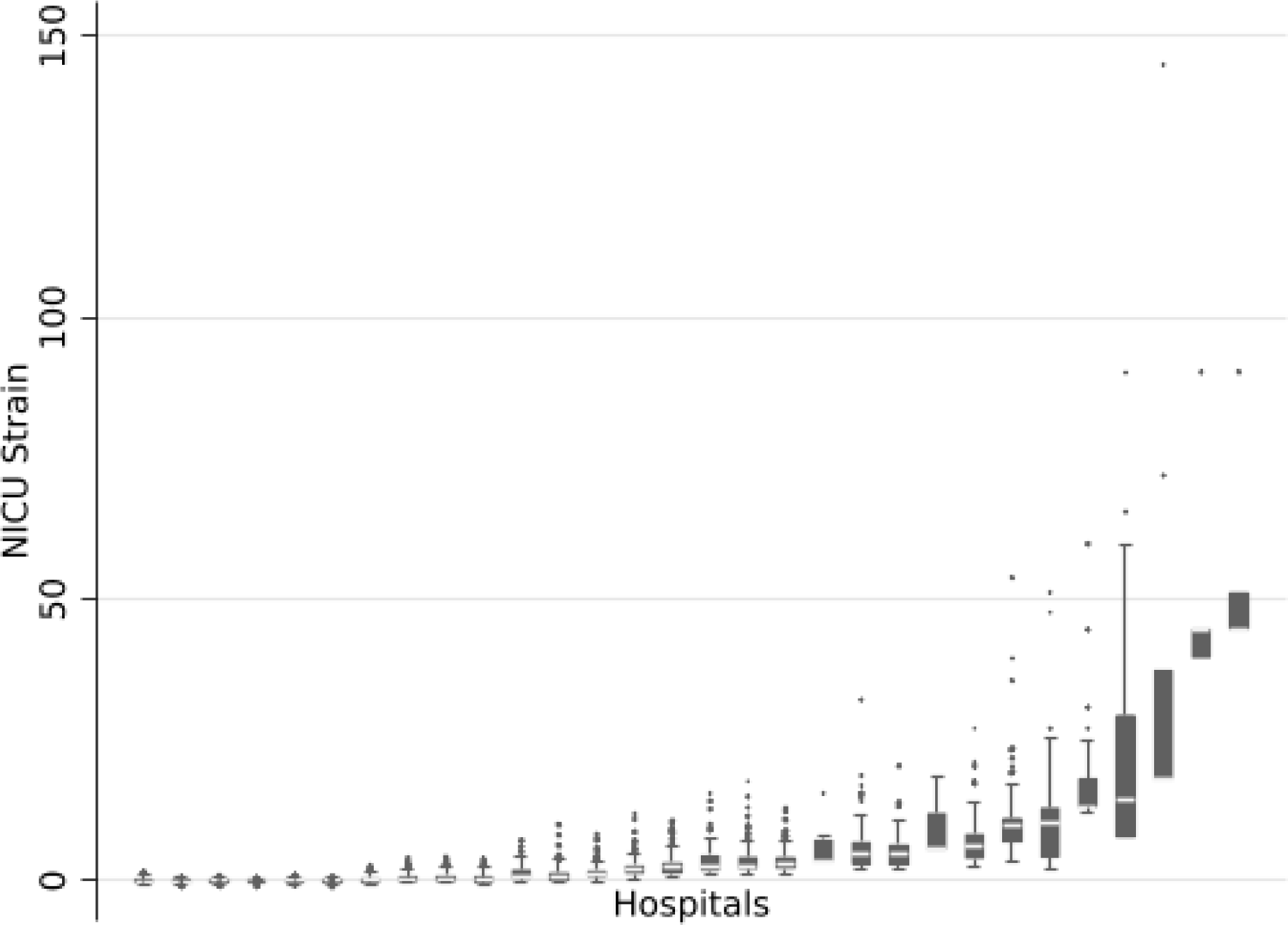
Variation in NICU strain by hospital. NICU strain was calculated as a standardized measure of the daily census of infants < 44 weeks GA with a congenital anomaly plus infants born < 33 weeks GA. Standardization was performed by calculating this daily census minus the average annual hospital census, divided by the average annual hospital census. Abbreviations: GA – Gestational age; NICU – Neonatal Intensive Care Unit.

**Figure 3.**
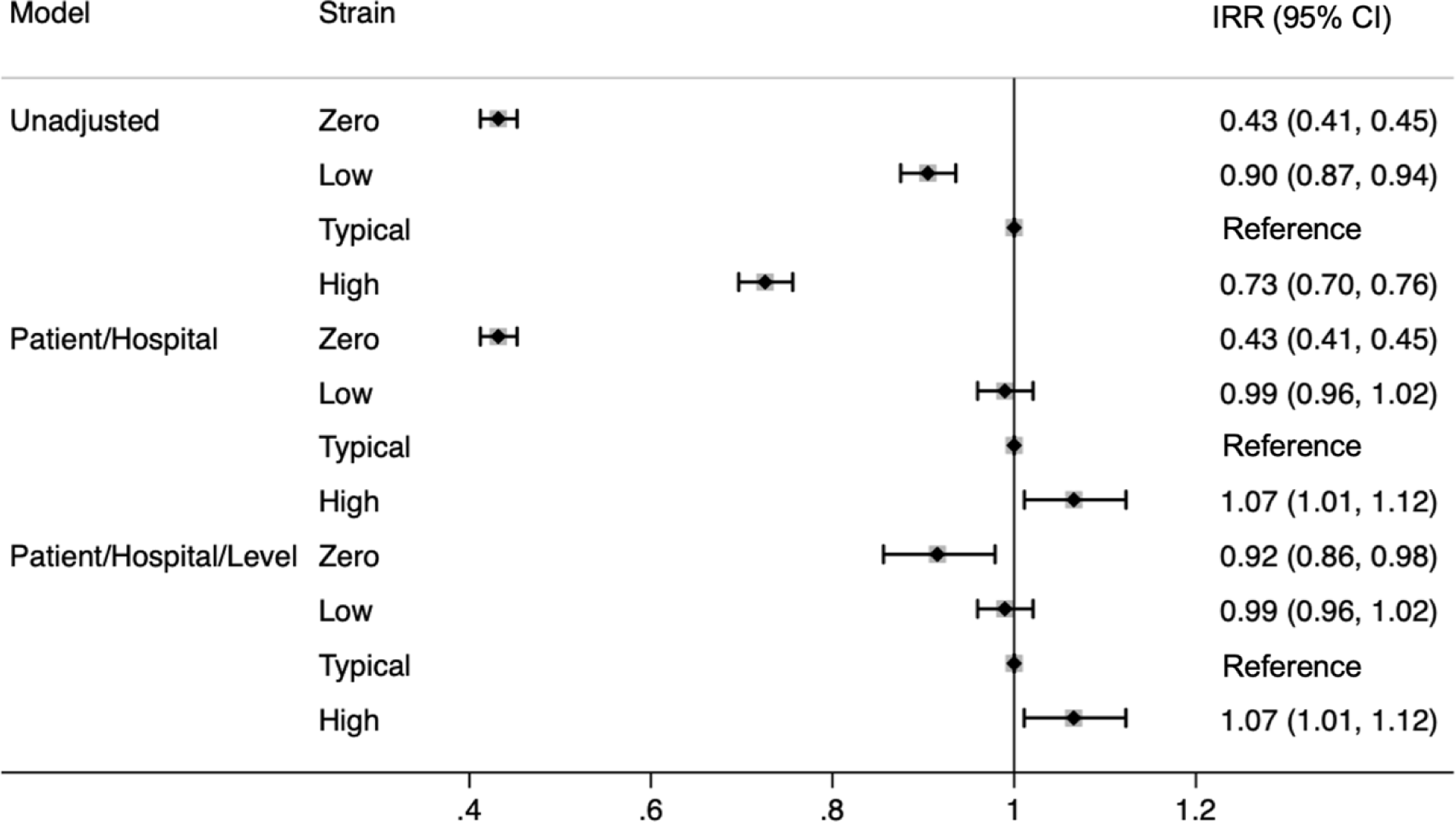
Association of NICU Strain with Composite Primary Outcome in Unadjusted and Adjusted Models. Models were multivariable modified poisson generalized linear mixed models. Patient characteristics include birthing parent variables (age, race and/or ethnicity, diabetes, hypertension, BMI, smoking, insurance, education, and cesarean section) and infant variables (gestational age, gender, small for gestational age, multiple gestation, multiple congenital anomalies). Hospital was controlled for with a fixed effect. AAP NICU level was included for the final set of models. Birth year was included in all models.

**Figure 4.**
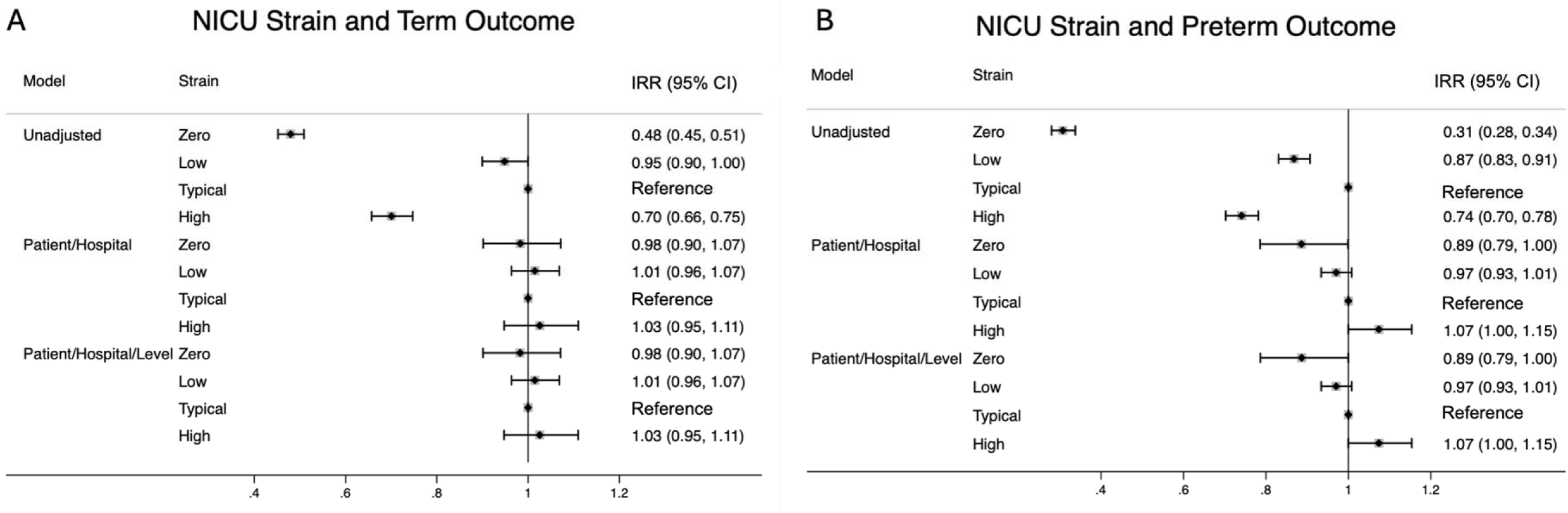
Association of NICU Strain with Preterm and Term Mortality and Morbidity in Unadjusted and Adjusted Models. Models were multivariable modified poisson generalized linear mixed models. Patient characteristics include birthing parent variables (age, race and/or ethnicity, diabetes, hypertension, BMI, smoking, insurance, education, and cesarean section) and infant variables (gestational age, gender, small for gestational age, multiple gestation, multiple congenital anomalies). Hospital was controlled for with a fixed effect. AAP NICU level was included for the final set of models. Birth year was included in all models.

### Sensitivity Analyses

We performed several sensitivity analyses to ensure the robustness of our findings. We examined the number of NICU admissions as an alternative measure of NICU strain and did not find an association with overall, term, or preterm outcomes (Supplemental Figures 2 and 3). We also excluded level 2 units and patients with zero values for strain and found consistent results (Supplemental Figures 4 and 5). Finally, we employed an alternate measure of standardization, dividing by the 95% value of annual hospital census, with consistent results.

## Results

The study cohort included 64,647 infants from 30 hospitals (273 hospital-years). There were 29,605 term (46%) and 25,042 preterm (54%) infants. In the cohort, 15,700 infants (24%) were exposed to zero strain; 16,396 (25%) experienced low strain; 20,552 (31%) experienced typical strain; and 11,999 (19%) experienced high strain at admission (Table 1). Infants exposed to typical strain were more frequently preterm, multiple gestation, and had a major congenital anomaly. Infants exposed to typical strain at admission were more often born to Black birthing parents with government insurance, diabetes, hypertension, and obesity. These infants were more often delivered via cesarean section (Table 1). Infants born during a period of typical strain were also more likely to be born at a non-profit, level 4 hospital with high annual birth volume (Supplemental Table 2).

**Table 1.**
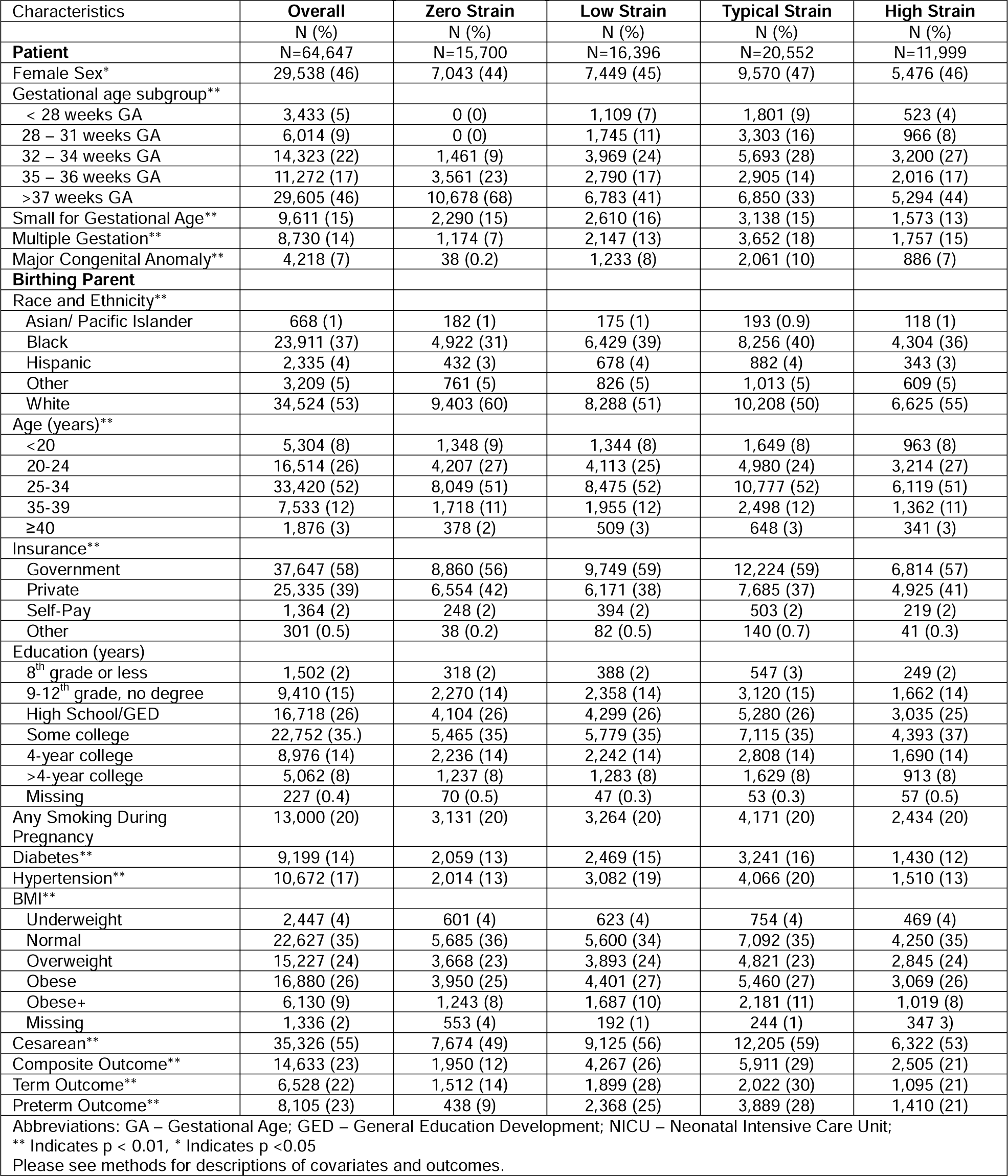
Perinatal Characteristics Overall and by NICU Strain Categories.

Figure 2 depicts the within and between variability of admission NICU strain by hospital. Hospitals with average daily hospital census of infants < 44 weeks GA with a congenital anomaly plus all infants born < 33 weeks GA less than 1 demonstrated more variability in NICU strain as the measure was standardized by dividing by this value, which is less than 1. Including a hospital fixed effect in our models allows us to interpret the strain variable as change of strain within a given hospital, allowing us to interpret our findings in the midst of this hospital level variability.

In unadjusted bivariate analyses, infants born during times of typical strain for a given hospital more frequently experienced composite, term, and preterm adverse outcomes (29% in the typical strain group vs. 21% in the high strain group and 12% in the zero strain group; Table 1). In unadjusted analyses, there was a decreased incidence rate ratio of the primary outcome in infants exposed to zero strain (IRR 0.43, 95% CI 0.41 – 0.45), low strain (IRR 0.90, 95% CI 0.87 – 0.94), and high strain (IRR 0.73, 95% CI 0.70 – 0.76) compared to typical strain.

In models adjusted for patient covariates with a fixed effect for hospital, exposure to birth during a period of high NICU strain for a given hospital was associated with an increased risk of mortality and morbidity (aIRR 1.07, 95% CI 1.01-1.12) compared to birth during typical strain.

This suggests that within a single hospital, birth during increased NICU strain was associated with increased mortality and morbidity when controlling for patient covariates. In adjusted models, birth during a period of zero strain was associated with a decreased risk of mortality and morbidity (aIRR 0.43, 95% CI 0.41-0.45) compared to birth during typical strain. When we used a fixed hospital effect with neonatal level of care, these associations persisted for both birth during times of either high or zero NICU strain (high strain aIRR 1.07, 95% CI 1.01-1.12; zero strain aIRR 0.92, 95% CI 0.86-0.98). Neonatal level of care did not have a significant impact on outcomes (level 3 aIRR 0.81, 95% CI 0.59-1.11) in these models.

In models examining secondary outcomes unadjusted for covariates, birth during both zero and high strain were associated with a lower relative risk of term outcomes (zero strain IRR 0.48, 95% CI 0.45 – 0.51; high strain RR 0.70, 95% CI 0.66 – 0.75) and preterm outcomes (zero strain IRR 0.31, 95% CI 0.28 – 0.34; high strain RR 0.74, 95% CI 0.70 – 0.78). After adjustment for patient and hospital covariates, zero strain remained associated with a decreased risk of the preterm outcomes (aIRR 0.89, 95% CI 0.79-0.99).

## Discussion

Research in the adult ICU demonstrated association with ICU capacity strain and patient outcomes, such as mortality, and quality of care.^11,13–16^ This study examines the association of NICU strain at admission with neonatal mortality and morbidity. To do this, we developed a novel measure of NICU strain, capturing both census and patient risk at admission, and standardized for the typical daily census of high-risk infants experienced within that NICU for a given year. In adjusted models for patient and hospital characteristics, exposure to high NICU strain on admission was associated with increased risk of morbidity and mortality.

This study is the first to report the association with NICU strain at admission with neonatal mortality and morbidity, consistent with findings in the adult ICU literature. Unlike the adult ICU however, the NICU contains both a mix of critically ill and convalescing patients awaiting appropriate development of feeding, breathing, and thermoregulation. This leads to NICUs have a longer average length of stay (approximately 13 days)^44^ compared to an adult ICU (approximately 3 days).^45^ It is particularly noteworthy that NICU strain at admission is associated with adverse neonatal outcomes because the NICU population is exposed to a longer admission period with varying exposure to strain. Our work supports prior literature highlighting the importance of the admission period surround delivery as essential in shaping neonatal outcomes.^5^

Increased adult ICU strain, measured through acuity-adjusted census and admissions, is associated with in-hospital adult mortality and decreased safety practices, including routine prophylaxis for venous thromboembolism and stress ulcers and routine sepsis admission procedures.^11,13,15,16^ Interestingly, our study suggests that in the NICU, only patient risk-adjusted census is associated with increased neonatal mortality and morbidity. One potential hypothesis for this finding is that births, and thus NICU admissions, may occur at a more regular cadence and thus cause less stress to the NICU system compared to an unplanned critical care admission. Future work examining the influence of NICU strain should consider examining whether admissions represent a substantial enough stress to influence more subtle measures, including quality of care. Prior literature reporting increased risk of sepsis during periods of increased proportion of infants <32 weeks GA in the NICU suggest that more subtle stressors may have an influence on quality of care and outcomes in the NICU.^17^ Future work may also consider examining the association of NICU strain with discharge and transfer, an association that has been described in prior work, and with birthing parent outcomes.^18,46^

This study has limitations. First, this study reports the influence of NICU strain in a single state given the quantity of daily data. Further examination in additional states is necessary to ensure generalizability. Second, while we adjusted for patient and birthing parent covariates, including presence of congenital anomalies and gestational age, to capture patient acuity, our data source did not contain additional clinical variables, such as days on the ventilator, to further control for patient acuity. Future work should explore whether incorporation of additional variables beyond congenital anomalies and gestational age will allow for more accurate identification of a high risk and acuity population contributing to NICU strain. Finally, while strain categories were defined using distributions of NICU strain, further research to establish optimal thresholds for high NICU strain are merited.

Despite these limitations, this study has many strengths. It uses a large, multiyear dataset of linked birthing parent-infant data capturing a variety of hospital types to create a novel definition of NICU capacity strain. It is the first study to report an association between NICU strain and neonatal morbidity and mortality. Additionally, it controls for other key hospital-level drivers of outcomes through a fixed effect and incorporation of neonatal levels of care. Our work indicates that NICU strain is an important and separate driver of outcome variability. By providing a definition of NICU strain, this work offers a first look into better understanding how NICU capacity strain influences neonatal outcome variation and associated disparities.

Future work is needed to better understand how NICU capacity strain may be mitigated. Longitudinal studies demonstrate a 42% increase in NICU beds from 1991 to 2017 without a clear relationship to associated increased in newborn risk.^47^ National studies may consider how this increase in NICU bed supply has influenced NICU strain over time. Additionally, the observed rise in discretionary NICU admissions for larger and less premature infants seen in recent years may also lead to increased capacity strain in the NICU, potentially negatively impacting patient outcomes.^48,49^ Our study examined the influence of NICU strain on admission only, and further work is needed to evaluate how the influence of NICU strain changes throughout the hospitalization. Finally, prior research suggests that nurse-staffing ratios are highly relevant to the very preterm infant population.^50^ Further investigation into how resources such as nurse staffing influence the relationship between NICU strain and patient outcomes are warranted.

## Conclusion

Exposure to high NICU strain at admission was associated with increased risk of neonatal mortality and morbidity when adjusting for infant, birthing parent, and hospital characteristics, including level of care. Further research is merited to characterize the influence of NICU strain on care processes and quality of care, as well as the influence of changing NICU strain over the course of the hospitalization. Future studies are needed to understand if available hospital resources, such as nurse: patient ratios, can mitigate the adverse effects of high NICU strain. Ultimately, this work supports the importance of capacity management in the NICU to optimize patient outcomes.

## Contributors Statement Page

Dr. Elizabeth Salazar conceptualized and designed the study and analyses, drafted the initial manuscript, reviewed, and revised the manuscript.

Brielle Formanowski and Molly Passarella carried out the analyses, critically reviewed and revised the manuscript.

Dr. Scott Lorch conceptualized and designed the study, obtained grant funding for the creation of the data source for the project, as well as critically reviewed and revised the manuscript.

Dr. Ciaran Phibbs provided critical feedback on study methodology, obtained grant funding for the creation of the data source for the project, as well as reviewed and revised the manuscript. All authors approved the final manuscript as submitted and agree to be accountable for all aspects of the work.

Dr. Jeannette Rogowski, Dr. Erika Edwards, and Dr. Scott Halpern provided critical feedback on study methodology, reviewed, and revised the manuscript.

## Data Availability

All data produced in the present study are available should a data use agreement be reached with the source and a reasonable request made to the authors.

## Acknowledgements

**SC:** This information is from the records of the Revenue and Fiscal Affairs Office, Health and Demographics Section, South Carolina. Our authorization to release this information does not imply endorsement of this study or its findings by either the revenue and fiscal affairs office or the data oversight council.

The views expressed in this article are those of the authors and do not necessarily reflect the position or policy of the Department of Veterans Affairs or the United States government.

## Abbreviations

CLD: Chronic Lung Disease
GA: Gestational Age
IVH: Intraventricular hemorrhage
NEC: Necrotizing enterocolitis
NICU: Neonatal Intensive Care Unit
PMA: Post Menstrual Age
ROP: Retinopathy of prematurity
sIVH: Severe intraventricular hemorrhage
VLBW: Very low birth weight

## Works Cited

1. Phibbs C, Smith Hughes C, Schmitt S, Passarella M, Lorch S. Who’s in the NICU? A population-level analysis. Journal of Perinatology. 2024;In Press

2. Profit J, Gould JB, Bennett M, et al. The Association of Level of Care with NICU Quality. Pediatrics. 2016/03// 2016;137(3):e20144210. doi:10.1542/peds.2014-4210

3. Lapcharoensap W, Gage SC, Kan P, et al. Hospital variation and risk factors for bronchopulmonary dysplasia in a population-based cohort. JAMA pediatrics. 2015/02/01/ 2015;169(2):e143676. doi:10.1001/jamapediatrics.2014.3676

4. Horbar JD, Edwards EM, Greenberg LT, et al. Variation in performance of neonatal intensive care units in the United States. JAMA Pediatrics. 2017/03/01/ 2017;171(3):e164396-e164396. doi:10.1001/jamapediatrics.2016.4396

5. Lorch SA, Baiocchi M, Ahlberg C, Small D. The differential impact of delivery hospital on the outcomes of premature infants. Pediatrics. 2012 2012;130(2):270–278. doi:10.1542/PEDS.2011-2820

6. Phibbs CS, Baker LC, Caughey AB, Danielsen B, Schmitt SK, Phibbs RH. Level and volume of neonatal intensive care and mortality in very-low-birth-weight infants. New England Journal of Medicine. 2007 2007;356(21):2165–2175. doi:10.1056/NEJMsa065029

7. Jensen EA, Lorch SA. Effects of a Birth Hospital’s Neonatal Intensive Care Unit Level and Annual Volume of Very Low-Birth-Weight Infant Deliveries on Morbidity and Mortality. JAMA Pediatrics. 2015;169(8):e151906. doi:10.1001/jamapediatrics.2015.1906

8. Chung JH, Phibbs CS, Boscardin WJ, Kominski GF, Ortega AN, Needleman J. The effect of neonatal intensive care level and hospital volume on mortality of very low birth weight infants. Med Care. 2010 2010;48(7):635–644.

9. Halpern SD. ICU capacity strain and the quality and allocation of critical care. Current Opinion in Critical Care. 2011 2011;17(6):648–657. doi:10.1097/MCC.0b013e32834c7a53

10. Duggal A, Mathews KS. Impact of ICU Strain on Outcomes. Current Opinion in Critical Care. 2022 2022;28(6):667–673. doi:10.1097/mcc.0000000000000993

11. Gabler NB, Ratcliffe SJ, Wagner J, et al. Mortality among patients admitted to strained intensive care units. American Journal of Respiratory and Critical Care Medicine. 2013 2013;188(7):800–806. doi:10.1164/rccm.201304-0622OC

12. Brown SES, Ratcliffe SJ, Halpern SD. An empirical comparison of key statistical attributes among potential ICU quality indicators. Critical Care Medicine. 2014 2014;42(8):1821–1831. doi:10.1097/CCM.0000000000000334

13. Weissman GE, Gabler NB, Brown SES, Halpern SD. Intensive care unit capacity strain and adherence to prophylaxis guidelines. Journal of Critical Care. 2015 2015;30(6):1303–1309. doi:10.1016/j.jcrc.2015.08.015

14. Wilcox ME, Harrison DG, Patel AJ, Rowan KM. Higher ICU Capacity Strain Is Associated With Increased Acute Mortality in Closed ICUs. Critical Care Medicine. 2020 2020;48(5):709–716. doi:10.1097/ccm.0000000000004283

15. Anesi GL, Dress E, Chowdhury M, et al. Hospital Strain and Variation in Sepsis ICU Admission Practices and Associated Outcomes. Critical Care Explorations. 2023 2023;5(2):e0858. doi:10.1097/cce.0000000000000858

16. Bravata DM, Perkins AJ, Myers LJ, et al. Association of Intensive Care Unit Patient Load and Demand With Mortality Rates in US Department of Veterans Affairs Hospitals During the COVID-19 Pandemic. Jama Network Open. 2021 2021;4(4):1. doi:10.1001/jamanetworkopen.2020.34266

17. Goldstein ND, Eppes SC, Ingraham BC, Paul DA. Characteristics of late-onset sepsis in the NICU: Does occupancy impact risk of infection? Journal of Perinatology. 2016 2016;36(9):753–757. doi:10.1038/jp.2016.71

18. Profit J, McCormick MC, Escobar GJ, et al. Neonatal intensive care unit census influences discharge of moderately preterm infants. Pediatrics. 2007/02// 2007;119(2):314-319. doi:10.1542/peds.2005-2909

19. Herrchen B, Gould JB, Nesbitt TS. Vital statistics linked birth/infant death and hospital discharge record linkage for epidemiological studies. Comput Biomed Res. 1997 1997;30(4):290–305. doi:10.1006/cbmr.1997.1448

20. American Hospital Assocation. AHA Annual Survey. https://www.ahadata.com/aha-annual-survey-database

21. Jensen EA, Lorch SA. Effects of a Birth Hospital’s Neonatal Intensive Care Unit Level and Annual Volume of Very Low-Birth-Weight Infant Deliveries on Morbidity and Mortality. JAMA pediatrics. 2015/08/01/ 2015;169(8):e151906. doi:10.1001/jamapediatrics.2015.1906

22. von Elm E, Altman DG, Egger M, et al. The Strengthening the Reporting of Observational Studies in Epidemiology (STROBE) statement: guidelines for reporting observational studies. Ann Intern Med. 2007 2007;147(8):573–577. doi:10.7326/0003-4819-147-8-200710160-0001010.

23. Phibbs CS, Passarella M, Schmitt S, Rogowski J, Lorch SA. Understanding the Relative Contributions of Prematurity and Congenital Anomalies to Neonatal Mortality. Journal of Perinatology. 2022 2022;42(5):569–573. doi:10.1038/s41372-021-01298-x

24. Joshi NS, Flaherman VJ, Halpern-Felsher B, Chung EK, Congdon JL, Lee HC. Admission and Care Practices in United States Well Newborn Nurseries. 2023 2023;13(3):208–216. doi:10.1542/HPEDS.2022-006882

25. Cerullo M, Lin Y-L, Rauh-Hain JA, Ho V, Offodile Ii AC. Financial Impacts And Operational Implications Of Private Equity Acquisition Of US Hospitals. Health Affairs. 2022;41(4):523–530. doi:10.1377/hlthaff.2021.01284

26. Handley SC, Passarella M, Interrante JD, Kozhimannil KB, Lorch SA. Perinatal Outcomes for Rural Obstetric Patients and Neonates in Rural-Located and Metropolitan-Located Hospitals. Journal of Perinatology. 2022 2022;42(12):1600–1606. doi:10.1038/s41372-022-01490-7

27. California Maternal Quality Care C. Unexpected complications in term newborns. 2018.

28. Tyson JE, Parikh NA, Langer J, et al. Intensive care for extreme prematurity--moving beyond gestational age. The New England journal of medicine. 2008/04/17/ 2008;358(16):1672-1681. doi:10.1056/NEJMOA073059

29. Fenton TR. A new growth chart for preterm babies: Babson and Benda’s chart updated with recent data and a new format. BMC pediatrics. 2003/12/16/ 2003;3 doi:10.1186/1471-2431-3-13

30. Clausson B, Gardosi J, Francis A, Cnattingius S. Perinatal outcome in SGA births defined by customised versus population-based birthweight standards. BJOG. 2001 2001;108(8):830–834.

31. Howell EA, Hebert P, Chatterjee S, Kleinman LC, Chassin MR. Black/white differences in very low birth weight neonatal mortality rates among New York City hospitals. Pediatrics. 2008/03// 2008;121(3)doi:10.1542/PEDS.2007-0910

32. Cleary-Goldman J, Malone FD, Vidaver J, et al. Impact of maternal age on obstetric outcome. Obstetrics and gynecology. 2005 2005;105(5 Pt 1):983-990. doi:10.1097/01.AOG.0000158118.75532.51

33. Kozhimannil KB, Shippee TP, Adegoke O, Vemig BA. Trends in hospital-based childbirth care: the role of health insurance. Am J Manag Care. 2013 2013;19(4):e125–32.

34. Arntzen A, Samuelsen SO, Bakketeig LS, Stoltenberg C. Socioeconomic status and risk of infant death. A population-based study of trends in Norway, 1967–1998. Int J Epidemiol. 2004 2004;33(2):279-88.

35. Liu B, Xu G, Sun Y, et al. Maternal cigarette smoking before and during pregnancy and the risk of preterm birth: A dose–response analysis of 25 million mother–infant pairs. PLOS Medicine. 2020 2020;17(8):e1003158. doi:10.1371/JOURNAL.PMED.1003158

36. Casson IF, Clarke CA, Howard CV, et al. Outcomes of pregnancy in insulin dependent diabetic women: results of a five year population cohort study. Bmj. 1997 1997;315(7103):275-278. doi:10.1136/bmj.315.7103.275

37. McBride CA, Bernstein IM, Badger GJ, Horbar JD, Soll RF. The effect of maternal hypertension on mortality in infants 22-29 weeks gestation. Pregnancy Hypertens. 2005 2005;5(4):362–366.

38. Liu B, Xu G, Sun Y, et al. Association between maternal pre-pregnancy obesity and preterm birth according to maternal age and race or ethnicity: a population-based study. Lancet Diabetes Endocrinol. 2019 2019;7(9):707–714. doi:10.1016/S2213-8587(19)30193-7

39. Korb D, Goffinet F, Seco A, Chevret S, Deneux-Tharaux C. Risk of severe maternal morbidity associated with cesarean delivery and the role of maternal age: a population-based propensity score analysis. CMAJ. 2019 2019;191(13):E352–E360.

40. Stoll BJ, Hansen NI, Bell EF, et al. Trends in Care Practices, Morbidity, and Mortality of Extremely Preterm Neonates, 1993-2012. JAMA. 2015/09/08/ 2015;314(10):1039-1051. doi:10.1001/jama.2015.10244

41. Hillemeier MM, Weisman CS, Chase GA, Dyer AM. Individual and community predictors of preterm birth and low birthweight along the rural-urban continuum in central Pennsylvania. J Rural Health. 2007 2007;23(1):42–8.

42. Snowden JM, Cheng YW, Kontgis CP, Caughey AB. The association between hospital obstetric volume and perinatal outcomes in California. Am J Obstet Gynecol. 2012 2012;207(6):478.e1-7. doi:10.1016/j.ajog.2012.09.029

43. Stark AR, Pursley DM, Papile L-A, et al. Standards for Levels of Neonatal Care: II, III, and IV. Pediatrics. 2023 2023;151(6):e2023061957. doi:10.1542/peds.2023-061957

44. Dimes Mo. Special Care Nursery Admissions. https://www.kff.org/wp-content/uploads/sites/2/2013/01/nicu_summary_final.pdf.

45. Cline SD, Schertz RAK, Feucht EC. Expedited admission of patients decreases duration of mechanical ventilation and shortens ICU stay. The American Journal of Emergency Medicine. 2009;27(7):843–846. doi:10.1016/j.ajem.2008.04.018

46. Beltempo M, Patel S, Platt RW, et al. Association of nurse staffing and unit occupancy with mortality and morbidity among very preterm infants: a multicentre study. Archives of disease in childhood Fetal and neonatal edition. 2023/07// 2023;108(4):387-393. doi:10.1136/archdischild-2022-324414

47. Davis R, Stuchlik PM, Goodman DC. The Relationship Between Regional Growth in Neonatal Intensive Care Capacity and Perinatal Risk. Medical Care. 2023 2023;doi:10.1097/mlr.0000000000001893

48. Harrison W, Wasserman JR, Goodman DC. Regional Variation in Neonatal Intensive Care Admissions and the Relationship to Bed Supply. The Journal of Pediatrics. 2018 2018;doi:10.1016/j.jpeds.2017.08.028

49. Goodman DC, Ganduglia-Cazaban C, Franzini L, et al. Neonatal Intensive Care Variation in Medicaid-Insured Newborns: A Population-Based Study. The Journal of Pediatrics. 2019 2019;209:44–51.e2. doi:10.1016/j.jpeds.2019.02.014

50. Beltempo M, Lacroix G, Cabot M, Blais R, Piedboeuf B. Association of nursing overtime, nurse staffing and unit occupancy with medical incidents and outcomes of very preterm infants. Journal of Perinatology: Official Journal of the California Perinatal Association. 2018/02// 2018;38(2):175-180. doi:10.1038/jp.2017.146

