## Supplemental for "The Association of NICU Strain with Neonatal Mortality and Morbidity"

Supplemental Table 1: ICD-9 Codes for Outcome

| Supplemental Table 1. ICD-9-CM Codes for Defined Study Variables | | |
|---|---|---|
| Variables | ICM-9-CM Codes | ICD-10-CM Codes |
| Necrotizing Enterocolitis | 7775, 77750, 77751, 77752, 77753 | P771, P772, P773, P779 |
| Intraventricular Hemorrhage Grade 3 or 4 | 77213, 77214 | P5221, P5222 |
| Chronic Lung Disease | 7707 | P270, P271, P278, P279 |
| Retinopathy of Prematurity | 36220, 36221, 36223, 36224, 36225, 36226, 36227, 36229 | H35101, H35102, H35103, H35109, H35121, H35122, H35123, H35129, H35131, H35132, H35133, H35139, H35141, H35142, H35143, H35149, H35151, H35152, H35153, H35159, H35161, H35162, H35163, H35169, H35171, H35172, H35173, H35179, H3520, H3521, H3522, H3523 |
| Any hypertension | 64200, 64201, 64202, 64203, 64204, 64210, 64211, 64212, 64213, 64214, 64220, 64221, 64222, 64223, 64224, 64230, 64232, 64233, 64234, 64240, 64241, 64242, 64243, 64244, 64250, 64251, 64252, 64253, 64254, 64270, 64271, 64272, 64273, 64274 | I10, O100, O1001, O1002, O1003, O101, O1011, O1012, O1013, O102, O1021, O1022, O1023, O103, O1031, O1032, O1033, O104, O1041, O1042, O1043, O109, O1091, O1092, O1093<br>O111, O112, O114, O114, O115, O119<br>O131, O132, O133, O134, O135, O139 |
| Any diabetes | 6488, 64880, 64882, 64883, 64884, 250, 2500, 25000, 25001, 25002, 25003, 2501, 25010, 25011, 25012, 25013, 2502, 25020, 25021, 25022, 25023, 2503, 25030, 25031, 25032, 25033, 2504, 25040, 25041, 25042, 25043, 2505, 25050, 25051, 25052, 25053, 2506, 25060, 25061, 25062, 25063, 2507, 25070, 25071, 25072, 25073, 2508, 25080, 25081, 25082, 25083, 2509, 25090, 25091, 25092, 25093, 3572, 3602, 36201, 36641, 6480, 64800, 64801, 64802, 64803, 64804 | E080, E081, E082, E083, E084, E085, E086, E088, E089, E090, E091, E092, E093, E094, E095, E096, E098, E099, E101, E102, E103, E104, E105, E106, E108, E109,E110, E111, E112, E113, E114, E115, E116, E118, E119, E12, E130, E131, E132, E133, E134, E135, E136, E138, E139,O240, O241, O243, O244, O248, O249, Z794 |
| Any infection | 0031, 0312, 0362, 0380, 0381, 03810, 03811, 03812, 03819, 0382, 0383, 03840, 03841,<br>03842, 03843, 03844, 03849, 0388, 0389, 0545, 05472, 77181, 77183, 7907, 99932, 00321, 0360, 0361, 09040, 09042, 09049, 09489, 0949, 3200, 3201, 3202, 3203, 3207, 32081, 32082, 32089, 3209, 3229, 3240, 3241, 3249, 325, 326, 09941, 59000, 59010, 59011, 5902, 59080, 5909, 5950, 59589, 5959, 5990, 77182, 00322, 01180, 01190, 01194, 0310, 0339, 0391, 481, 4820, 4821, 4822, | A021, A327, A400, A401, A408, A409, A4101, A4102, A411, A412, A413, A414, A4150, A4151, A4152, A4153, A4159, A4181, A4189, A419, B377, P360, P3610, P3619, P362, P3630, P3639, P364, P365, P368, P369, P369, P372, R6520, R6521, R7881, T80211A, A5040, A5041, A5049, B1009, G000, G002, G003, G008, G009, G01, G02, G038, G039, G042, G060, G061, G062, A523, A5219, A170, N10, N110, N3000, N3080, N3090, N3091, P393, R8271, T83511A, T83518A, J22, A159, A420, J13, J14, J150, J151, J1520, J15211, J15212, J1529, J153, J154, J155, |

| | | |
|---|---|---|
| | 48230, 48231, 48232, 48239, 48240, 48241, 48242, 48249, 48281, 48282, 48283, 48289, 4829, 4830, 4831, 4838, 4843, 4848, 485, 486, 5130, 0010, 0030, 0040, 0051, 0052, 0053, 00589, 0059, 00800, 00804, 00809, 0082, 00841, 00842, 00843, 00845, 00846, 00847, 00849, 0085, 0090, 0091, 0092, 0392, 11285, 5695, 5582, 5670, 56723, 56729, 5678, 56789, 5679, 01402, 01404, 0022, 0023, 00320, 0038, 0039, 0041, 0048, 0049, 01000, 01890, 0208, 0209, 0239, 024, 0270, 0271, 0300, 0308, 0319, 03289, 0329, 0330, 0331, 0363, 03689, 0369, 037, 0398, 0399, 0400, 0403, 04041, 04082, 04089, 04100, 04101, 04102, 04103, 04104, 04105, 04109, 04110, 04111, 04112, 04119, 0412, 0413, 0414, 04141, 04149, 0415, 0416, 0417, 04181, 04182, 04183, 04184, 04185, 04186, 04189, 0419, 0739, 07888, 07988, 07998, 08881, 0900, 0901, 0902, 0905, 0907, 0909, 0919, 0920, 0929, 09389, 096, 0971, 0979, 0318, 0318, V091, V092, V093, V094, V0950, V0951, V096, V0970, V0980, V0981, V0990, V0991, V021, V022, V023, V0251, V0252, V0253, V0254, V0259, V027, V028, 7718, 77189, 9993, 99931, 99939, V1200, V1201, V1204, V1209, 0761, 0769, 0783, 01500, 01504, 01700, 03281, 03285, 0340, 035, 07798, 0903, 0913, 0955, 0980, 0982, 09840, 09849, 09886, 09889, 0990, 0993, 09950, 09954, 09959, 0998, 0999, 101, 1027, 37601, 38022, 38023, 38200, 3824, 3829, 38300, 3831, 3839, 4210, 42292, 42490, 42491, 42499, 449, 4510, 45111, 45119, 4512, 45181, 45182, 45183, 45184, 45189, 4519, 4572, 460, 4610, 4612, 4618, 4619, 462, 463, 4640, 46400, 46401, 46410, 46411, 46420, 46421, 46430, 46431, 5131, 51901, 5192, 5225, 5283, 56731, 5673, 5721, 5761, 6040, 6800, 6801, 6802, 6803, 6804, 6805, 6806, 6808, 68100, 68101, 68102, 68110, 68111, 6819, 6820, 6821, 6822, 6823, 6824, 6825, 6826, 6827, 6828, 6829, 683, 684, 6850, 6868, 6869, 71100, 71101, 71102, 71104, 71105, 71106, 71107, 71109, 71181, 71195, 72886, 73000, 73001, 73002, 73003, 73004, 73005, 73006, 73007, 73008, 73009, 73012, | J156, J157, J158, J159, J160, J168, J17, J180, J181, J182, J188, J189, J850, J851, J860, J869, J95851, P231, P232, P233, P234, P235, P236, P238, P239, A5004, A044, A045, A047, A0472, A048, A049, A088, A09, A421, K612, A020, K652, A5145, A028, A029, A219, A230, A2789, A305, A329, A33, A3700, A3790, A4289, A480, A483, A4851, A488, A4901, A4902, A491, A492, A493, A498, A499, A5008, A5009, A501, A502, A5059, A509, A5279, A530, A539, A7740, A78, B950, B951, B952, B953, B954, B955, B9561, B9562, B957, B958, B960, B961, B9620, B9622, B9629, B963, B964, B965, B966, B967, B9689, K3531, V091, V092, V093, B998, B999, P399, Z220, Z221, Z22321, Z22322, Z22330, Z22338, Z2239, Z224, Z228, Z229, A46, A5001, A5002, A5006, A5007, A5031, A5057, A510, A5274, A5277, A5409, A5430, A5431, A549, A568, A57, A671, A672, A740, A749, H00032, H00033, H6001, H6002, H6010, H6011, H6012, H66002, H66012, H6640, H6642, H6690, H6691, H6692, H6693, H7090, J340, J852, J853, J9851, K047, K6812, K6819, K830, L00, L0100, L0101, L0103, L0109, L0201, L0202, L0211, L02211, L02212, L02213, L02214, L02216, L02222, L0231, L0233, L02411, L02412, L02413, L02414, L02415, L02416, L02419, L02423, L02511, L02512, L02519, L02521, L02522, L02611, L02612, L02619, L02621, L02811, L02818, L02821, L0291, L0292, L03011, L03012, L03019, L03031, L03032, L03111, L03112, L03113, L03114, L03115, L03116, L03119, L03211, L03213, L03221, L03311, L03312, L03313, L03314, L03315, L03316, L03317, L03319, L03811, L03818, L0390, L040, L042, L043, L0592, L081, L0882, M726, M8600, M86021, M86051, M86052, M86061, M86062, M8610, M86112, M86121, M86122, M86132, M86151, M86152, M86172, M8618, M8619, M86621, M86651, M8668, M868X0, M868X1, M868X2, M868X3, M868X5, M868X6, M868X8, M868X9, M869, P381, P389, P390, P391, T847XXA, T8483XA, T8571XA, T85, 30A, T8579XA, G08, H05011, H10021, H10023, H10029, H1030, H1031, H1032, H1033, H44001, H44002, H44003, H44009, H6090, H6091, H6092, H6093, I301, I330, I38, I8001, I8002, I8011, I8012, I80201, I80211, I80233, I80291, I80299, I803, I808, I809, I891, K113, K122, K352, K353, K3532, |

| | | |
|---|---|---|
| | 73015, 73016, 73018, 73020, 73021, 73022, 73023, 73024, 73025, 73026, 73027, 73028, 73029, 73030, 73032, 73033, 73035, 73036, 73037, 73038, 73039, 73079, 73081, 73085, 73086, 73088, 73089, 73098, 7713, 7714, 7715, 7716, 99933, 2893, 36000, 36001, 36002, 36003, 36019, 36313, 36320, 37040, 37049, 3708, 3709, 37200, 37300, 3735, 37530, 47871, 5400, 5401, 5409, 541, 542, 7280, 72930, 72939, 7294, 37532, 37533, 38010, 38012, 38013, 38201, 13100, 13101, 4650, 1320, 1321, 1329, 0279, 0070, 0072, 0074, 0838, 0839, 0844, 0846, 08882, 1216, 1218, 1229, 1256, 1259, 1269, 1289, 1302, 1308, 1309, 1318, 1319, 1368, 1369, 1330, 1341, 46450, 4658, 4659, V0260, V0261, V0262, V0269, 04511, 07030, 07031, 07032, 07051, 07054, 07059, 07070, 0709, 5731, 5732, 3220, 0470, 0471, 0478, 0479, 048, 0490, 0491, 0498, 0499, 0530, 05319, 0543, 05829, 0621, 48881, 4800, 4801, 4802, 4808, 4809, 4841, 4870, 00861, 00862, 00863, 00864, 00866, 00867, 00869, 0088, 042, 04500, 04591, 0502, 0527, 0528, 0529, 0539, 05479, 0548, 0549, 0569, 0570, 0578, 0579, 05810, 05889, 05919, 0600, 064, 0663, 0701, 07423, 0743, 0748, 075, 0773, 0785, 07889, 0790, 0791, 0792, 0793, 0794, 07950, 07951, 07953, 07959, 0796, 07981, 07983, 07989, 07999, 4660, 4661, 46611, 46619, 4871, 4878, 488, 4880, 48802, 4881, 48811, 48812, 48882, 7710, 7711, 7712, 05320, 05329, 0540, 05410, 05413, 05419, 0542, 05440, 05441, 05442, 05443, 05449, 05473, 0740, 0778, 07799, 0780, 07810, 07811, 07812, 07819, 464, 11281, 11289, 1129, 1124, 3210, 3210, 11283, 1122, 4846, 4847, 7717, 1125, 1140, 1143, 1149, 11509, 1172, 1173, 1175, 1177, 1179, 118, 1363, 1100, 1101, 1102, 1103, 1104, 1105, 1108, 1109, 1110, 1111, 1118, 1119, 1120, 1121, 1123, 11282, 11284, 71168 32301, 32341, 3238, 32381, 3239 | K3580, K36, M00011, M00012, M00051, M00052, M00061, M00071, M00072, M00822, M00861, M009, M60003, M60051, M60852, M60862, M60872, M6088, M609, M61141, M65161, N4822, N610, I400, J0190, J020, J029, J0390, J040, J0410, J0411, J042, J0430, J0431, J050, J0510, J060, P394, P398, B86, A598, A599, B589, B59, B662, B80, B832, B851, B852, B880, B948, P371, P373, P378, P379, A8030, B179, B181, B182, B1910, B1920, B199, B251, B941, G030, A850, A858, A86, A870, A872, A878, A879, A888, B003, B004, T85738A, B250, J1000, J1001, J1008, J120, J121, J122, J123, J1289, J129, P230, A080, A0811, A0819, A082, A0832, A0839, A084, A925, A928, B007, B0089, B009, B019, B029, B069, B083, B1081, B20, B258, B259, B269, B2709, B3322, B334, B338, B341, B342, B348, B349, B970, B9710, B9711, B9712, B9719, B9729, B974, B9781, B9789, J101, J1089, J111, J201, J205, J206, J208, J209, J210, J211, J218, J219, P350, P351, P352, P353, P358, P359, A880, J09X2, A6002, A601, A630, B001, B0052, B0053, B0059, B078, B079, B081, B084, B085, B088, B09, B300, B303, B309, J00, L130, J069, B3789, B379, B464, B375, B3749, B371, B441, B3781, B376, B447, B4489, B449, B465, B488, B49, P375, B350, B351, B352, B354, B356, B358, B359, B368, B369, B370, B372, B373, B383, B463, G0481, G0490, G053, J22, T80218A, T80219A, T80212A |

| Supplemental Table 2. Perinatal Characteristics Overall and by NICU Strain Categories | | | | | |
|---|---|---|---|---|---|
| Characteristics | **Overall** | **Zero Strain** | **Low Strain** | **Typical Strain** | **High Strain** |
| | N (%) | N (%) | N (%) | N (%) | N (%) |
| **Hospital** | N=64,647 | N=15,700 | N=16,396 | N=20,552 | N=11,999 |
| Hospital Ownership** | | | | | |
| Government | 13,861 (21) | 6,615 (42) | 1,739 (11) | 1,373 (7) | 4,134 (34) |
| Non-Profit | 32,390 (50) | 7,242 (46) | 8,555 (52) | 11,469 (56) | 5,124 (43) |
| Profit | 18,396 (28) | 1,843 (12) | 6,102 (37) | 7,710 (38) | 2,741 (23) |
| Hospital Urban Influence Code** | | | | | |
| Metropolitan | 62,101 (96) | 15,368 (99) | 15,544 (95) | 19,913 (97) | 11,276 (94) |
| Micropolitan | 2,501 (4) | 303 (2) | 852 (5) | 639 (3) | 707 (6) |
| Noncore | 45 (0.1) | 29 (0.2) | 0 (0.0) | 0 (0) | 16 (0.1) |
| NICU Beds** | 34.00 (27) | 9.13 (5) | 47.92 (24) | 52.96 (21) | 13.94 (12) |
| NICU Level of Care** | | | | | |
| 2 | 8,036 (12) | 5,547 (35) | 0 (0) | 39 (0.2) | 2,450 (20) |
| 3 | 43,198 (67) | 10,153 (65) | 11,329 (69) | 12,269 (60) | 9,447 (79) |
| 4 | 13,413 (21) | 0 (0) | 5,067 (31) | 8,244 (40) | 102 (0.9) |
| Delivery Volume Category** | | | | | |
| 10-500 | 356 (0.6) | 227 (1) | 0 (0) | 0 (0) | 129 (1) |
| 501-1000 | 4,975 (7.7) | 3,346 (21) | 12 (0.1) | 86 (0.4) | 1,531 (13) |
| 1001-2000 | 17,467 (2) | 8,433 (54) | 1,901 (12) | 2,053 (10) | 5,080 (42) |
| >2000 | 41,849 (65) | 3,694 (24) | 14,483 (88) | 18,413 (90) | 5,259 (44) |

Abbreviations: GA – Gestational Age; GED – General Education Development; NICU – Neonatal Intensive Care Unit;
** Indicates p < 0.01, * Indicates p <0.05
Rurality is defined using Urban Influence Codes (UICs). UICs 1 or 2 are metropolitan. UICs 3, 5, or 8 are micropolitan. UICs 4, 6, 7, 9, 10, 11 or 12 are noncore.

Supplemental Figure 1. Consort Flow Diagram

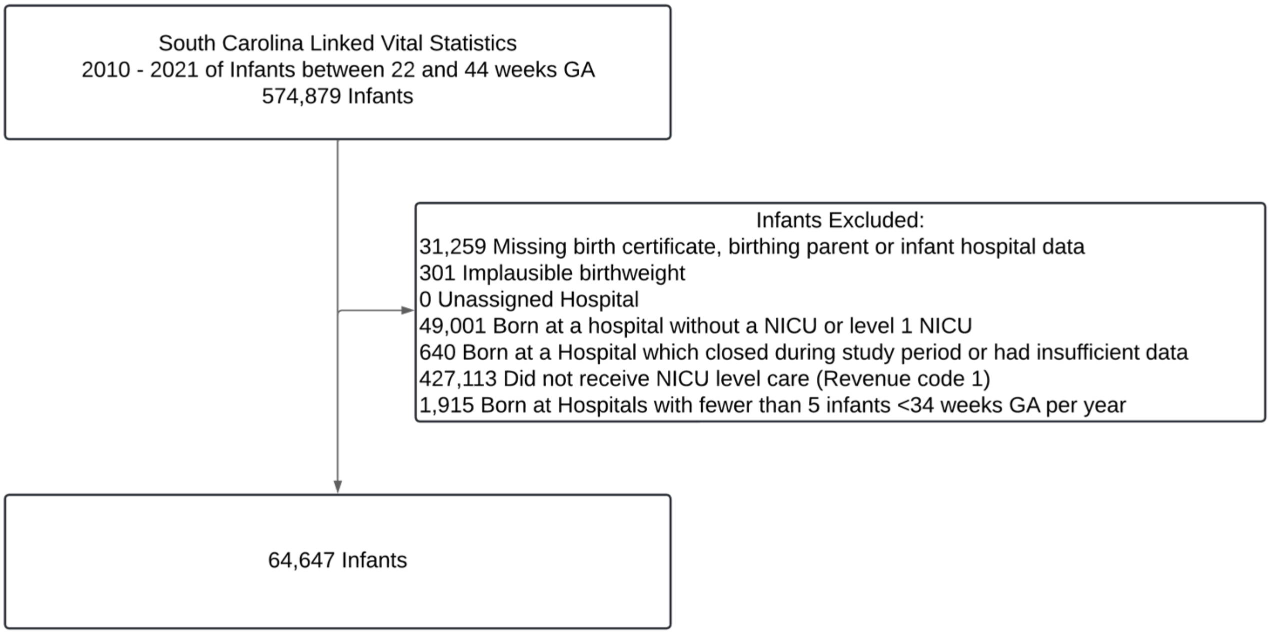


Supplemental Figure 2. NICU Admissions and Composite Outcome

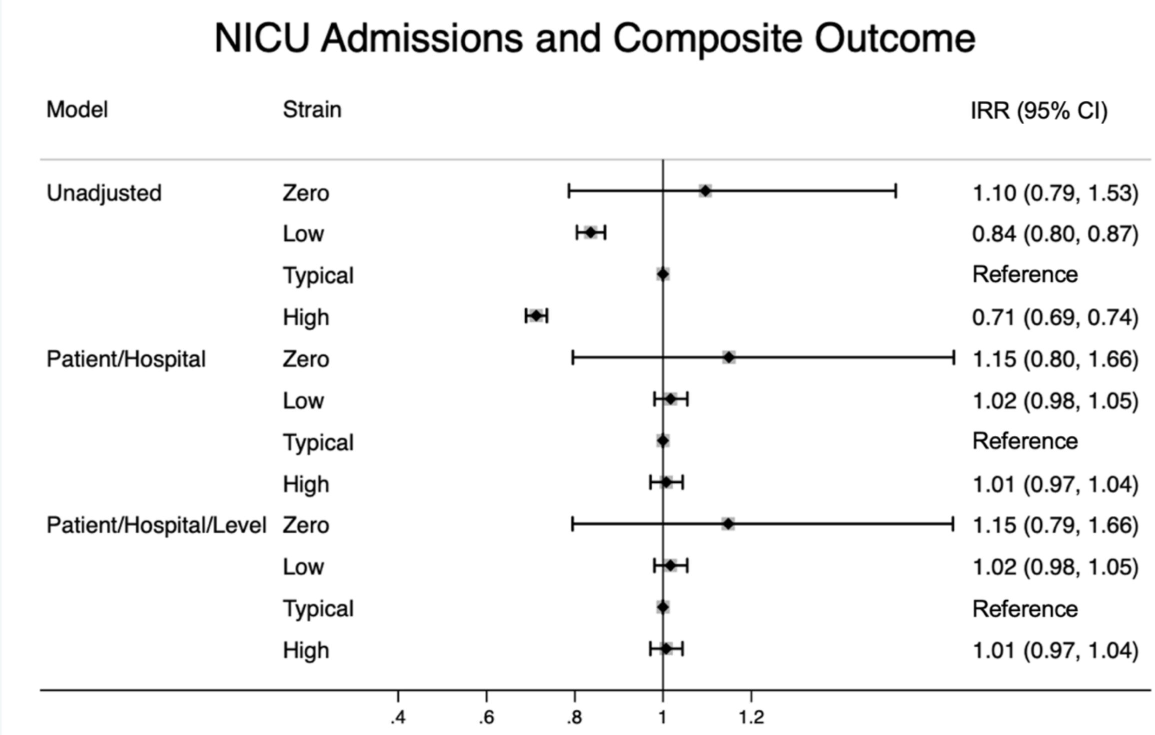


Models were multivariable modified poisson generalized linear mixed models. Patient characteristics include birthing parent variables (age, race and/or ethnicity, diabetes, hypertension, BMI, smoking, insurance, education, and cesarean section) and infant variables (gestational age, gender, small for gestational age, multiple gestation, multiple congenital anomalies). Hospital was controlled for with a fixed effect. AAP NICU level was included for the final set of models. Birth year was included in all models.

Supplemental Figure 3. NICU Admissions and Term and Preterm Outcomes

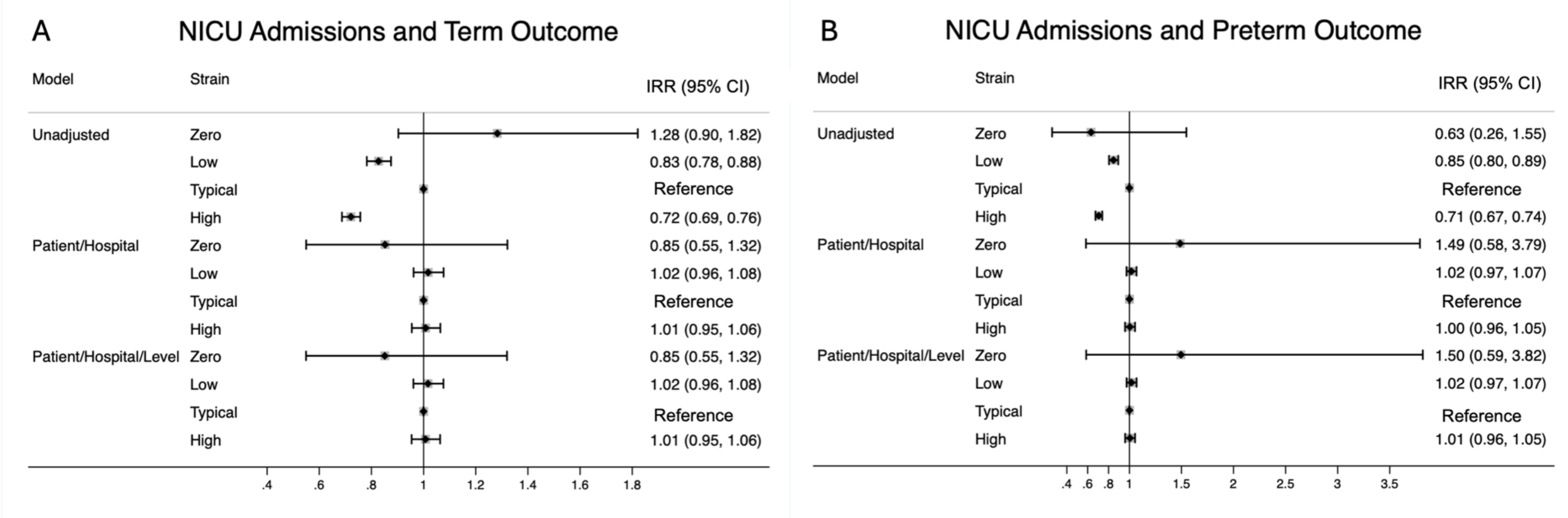


Models were multivariable modified poisson generalized linear mixed models. Patient characteristics include birthing parent variables (age, race and/or ethnicity, diabetes, hypertension, BMI, smoking, insurance, education, and cesarean section) and infant variables (gestational age, gender, small for gestational age, multiple gestation, multiple congenital anomalies). Hospital was controlled for with a fixed effect. AAP NICU level was included for the final set of models. Birth year was included in all models.

Supplemental Figure 4. NICU Strain and Composite Outcome with Level 2 Units Excluded

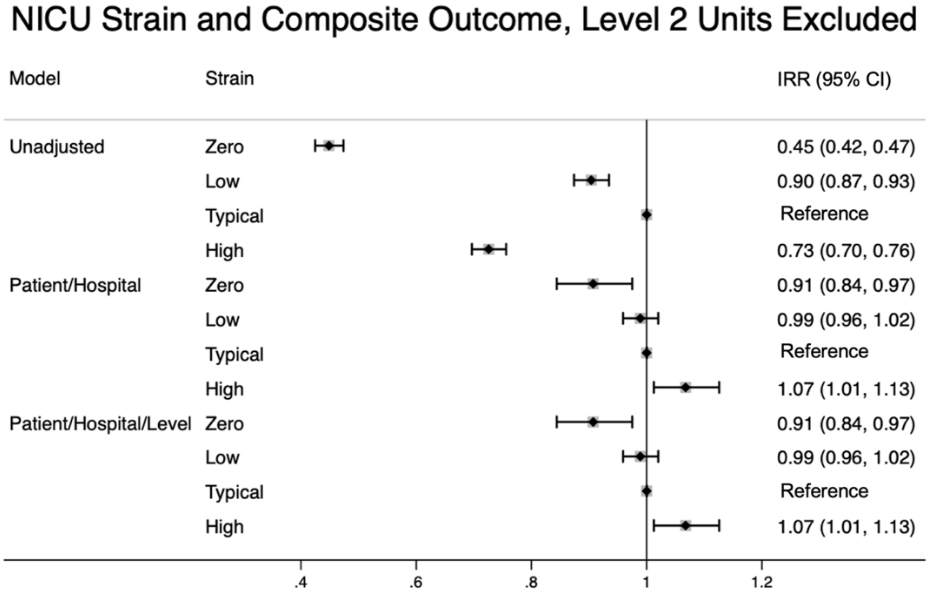


Models were multivariable modified poisson generalized linear mixed models. Patient characteristics include birthing parent variables (age, race and/or ethnicity, diabetes, hypertension, BMI, smoking, insurance, education, and cesarean section) and infant variables (gestational age, gender, small for gestational age, multiple gestation, multiple congenital anomalies). Hospital was controlled for with a fixed effect. AAP NICU level was included for the final set of models. Birth year was included in all models.

Supplemental Figure 5. NICU Strain and Composite Outcome with Zero Strain Values Excluded

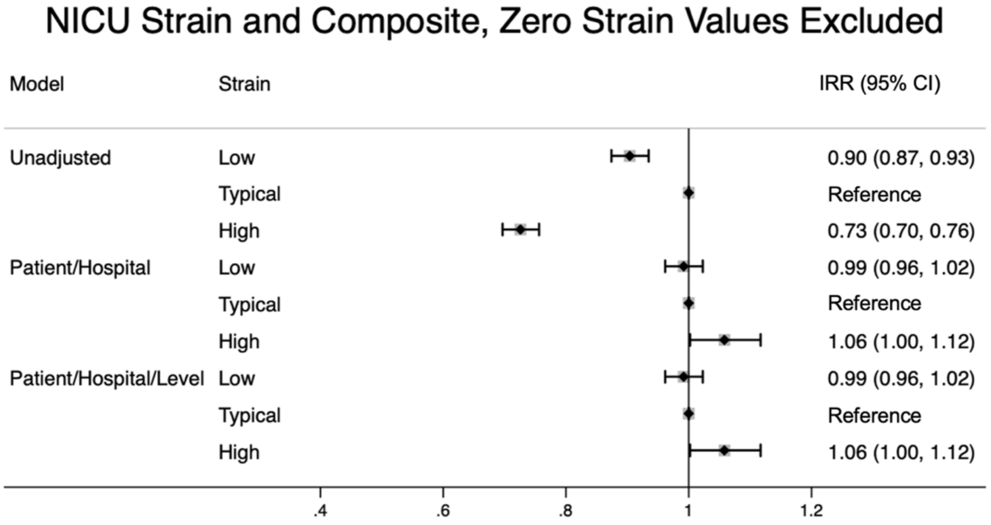


Models were multivariable modified poisson generalized linear mixed models. Patient characteristics include birthing parent variables (age, race and/or ethnicity, diabetes, hypertension, BMI, smoking, insurance, education, and cesarean section) and infant variables (gestational age, gender, small for gestational age, multiple gestation, multiple congenital anomalies). Hospital was controlled for with a fixed effect. AAP NICU level was included for the final set of models. Birth year was included in all models.
